# Why COVID-19 is not so spread in Africa: How does Ivermectin affect it?

**DOI:** 10.1101/2021.03.26.21254377

**Authors:** Hisaya Tanioka, Sayaka Tanioka, Kimitaka Kaga

**Affiliations:** Tanioka Clinic, Bunkyo-Ku Tokyo, Japan; National Institute of Sensory Organs, Tokyo Medical Center, Tokyo, 152-8902, Japan

**Keywords:** COVID-19, SARS-CoV-2, Africa, Ivermectin, Onchocerciasis

## Abstract

**Background:** Scientists have so far been unable to determine the reason for the low number of COVID-19 cases in Africa.

**Objective:** To evaluate the impact of ivermectin interventions for onchocerciasis on the morbidity, mortality, recovery, and fatality rates caused by COVID-19.

**Method:** A retrospective statistical analysis study of the impact of ivermectin against COVID-19 between the 31 onchocerciasis-endemic countries using the community-directed treatment with ivermectin (CDTI) and the non-endemic 22 countries in Africa. The morbidity, mortality, recovery rate, and fatality rate caused by COVID-19 were calculated from the WHO situation report in Africa. We investigated the onchocerciasis endemic 31 countries and the non-endemic 22 countries. Statistical comparisons used by the Welch test of them in the two groups were made.

**Results:** The morbidity and mortality were statistically significantly less in the 31 countries using CDTI. The recovery and fatality rates were not statistically significant difference. The average life expectancy was statistically significantly higher in the non-endemic countries.

**Conclusions:** The morbidity and mortality in the onchocerciasis endemic countries are lesser than those in the non-endemic ones. The community-directed onchocerciasis treatment with ivermectin is the most reasonable explanation for the decrease in morbidity and fatality rate in Africa. In areas where ivermectin is distributed to and used by the entire population, it leads to a significant reduction in mortality.

## Introduction

At least for now, it seems that Africa will be in completely different situations under the coronavirus infections. Some scientists have cited a higher proportion of young people [1,2], a warmer climate [3], and widespread BCG vaccination [4] as possible factors. While these are positive theories, they do not provide scientific evidence to explain why the spread of new coronavirus infections in Africa appears to be at a slower pace than in other parts of the world.

In the meantime, based on a growing data of recently reported data on a large number of published and unpublished trials, it is suggested that ivermectin being a well-known antiparasitic agent with antiviral activity and anti-inflammatory effects, has activity against SARS-CoV-2 [5]. On the other hand, ivermectin has been administered in Africa for onchocerciasis under the WHO strategy. In 2012, WHO’s neglected tropical diseases (NTD) Roadmap set a goal of elimination where feasible by 2020, and the African Programme for Onchocerciasis Control advanced the goal to elimination in 80% of countries by 2025 [6]. The community-directed treatment with ivermectin (CDTI) is the basic strategy to eradicate onchocerciasis in Africa. More than 99% of the infections have occurred in the 31 countries in Sub-Saharan Africa listed below: Angola, Benin, Burkina Faso, Burundi, Cameroon, Central Africa, Chad, Republic of Congo, Cote d’Ivoire, Democratic Republic of Congo, Equatorial Guinea, Ethiopia, Gabon, Ghana, Guinea, Guinea-Bissau, Kenya, Liberia, Malawi, Mali, Mozambique, Niger Nigeria, Rwanda, Senegal, Sierra Leone, South Sudan, Sudan, Togo, Uganda, Tanzania. In the rural populations of sub-Saharan Africa where health systems are weak and under-resourced, the community-directed treatment strategy is proving to be one of Africa’s most successful in reducing disease at low cost [7].

If ivermectin has an antiviral effect on SARS-CoV-2, the morbidity, mortality, recovery, and fatality rates caused by COVID-19 would be reduced in the community-directed treatment with ivermectin (CDTI) countries compared to non-endemic untreated ones. Therefore, epidemiological analyzes of the two groups are necessary. These results will validate the effect of ivermectin intervention on COVID-19. This study aims to evaluate the impact of ivermectin interventions for onchocerciasis on morbidity, mortality, recovery rate, and fatality rate caused by COVID-19.

## Materials

We divided into two group countries. One was 31 onchocerciasis-endemic countries (**Ivermectin group**) using the community-directed treatment with ivermectin (CDTI), and the other was 22 non-endemic countries (**non-Ivermectin group**). Each population and average life expectancy were obtained from the WHO Africa statics in 2019 [8]. The COVID-19 data was obtained from the WHO coronavirus disease Dashboard on January 15, 2021, and the COVID-19 Situation update for the WHO African Region [9]. Each morbidity, mortality, recovery rate, and fatality rate caused by COVID-19 were calculated from the collected data. **Table 1** showed the population (in millions), morbidity (number of cases and per million population), mortality (number of deaths and per million population), recovery rate (number of cases and percentage), and fatality rate (percentage) in the onchocerciasis-endemic 31 countries. **Table 2** showed them in the non-endemic 22 countries.

**Table 1.** Ivermectin 31 countries.

| Country | Pop<br>(M) | Morbidity<br>(N/M) | Mortality<br>(N/M) | Recovery<br>(N/%) | Fatality<br>(%) | Life<br>(y/o) |
| --- | --- | --- | --- | --- | --- | --- |
| Angola | 32.9 | 18765 / 571.0 | 431/13.1 | 16225 / 86.5 | 2.3 | 60.4 |
| Bénin | 12.1 | 3413 / 281.5 | 46 / 3.8 | 3245 / 95.1 | 1.3 | 61.2 |
| Buikna Fasso | 20.9 | 9000 / 430.6 | 101 / 4.8 | 7102 / 78.9 | 1.1 | 60.3 |
| Burundi | 11.9 | 1185 / 99.7 | 2 / 0.2 | 773 / 65.2 | 0.2 | 50.9 |
| Cameroon | 26.5 | 28010/1055.2 | 455/17.1 | 26861/95.9 | 1.6 | 58.5 |
| Cent Africa Re | 4.9 | 4973 / 1007.1 | 63 / 2.8 | 3181 / 64.0 | 1.3 | 52.2 |
| Chad | 16.4 | 2855 / 173.8 | 111 /6.8 | 2107 / 73.8 | 3.9 | 53.7 |
| De.Re.Congo | 84.1 | 20693 / 246.1 | 630 / 7.5 | 14804 / 71.5 | 3.0 | 60.0 |
| Rep. Congo | 5.5 | 7709 /1397.1 | 114 / 20.7 | 5846 / 75.8 | 1.5 | 64.5 |
| Côte d'Ivoire | 26.4 | 24856 / 942.3 | 141/5.3 | 23201/93.3 | 0.6 | 57.4 |
| Ecuat Guinea | 1.7 | 5356 / 3150.6 | 86 / 50.6 | 5189 / 96.9 | 1.6 | 58.1 |
| Ethiopia | 115.0 | 130772 / 1137.1 | 2029 / 17.7 | 116045 / 88.7 | 1.5 | 71.8 |
| Gabon | 2.2 | 9899 / 4499.5 | 66/ 30.0 | 9,658 / 97.6 | 0.7 | 65.8 |
| Ghana | 31.1 | 56981/ 1832.2 | 341/ 11.0 | 55236 / 96.9 | 0.7 | 63.5 |
| Guinea | 13.1 | 14098 / 1076.2 | 81/ 6.2 | 13320 / 94.5 | 0.6 | 60.7 |
| Guinea-Bissau | 2.0 | 2487 / 1243.5 | 45 / 22.5 | 2400 / 96.5 | 1.8 | 57.7 |
| Kenya | 53.8 | 99082 / 1841.7 | 2831/ 52.6 | 83324 / 84.0 | 2.9 | 65.8 |
| Liberia | 5.1 | 1887 / 370.0 | 84 / 16.5 | 1701 / 90.1 | 4.5 | 63.7 |
| Malawi | 19.1 | 11785 / 617.0 | 300 / 15.7 | 5992 / 50.8 | 2.5 | 63.8 |
| Mali | 20.3 | 7823 / 385.4 | 308 / 15.2 | 5531/ 70.7 | 3.9 | 58.5 |
| Mozambique | 31.3 | 25862 / 826.3 | 234 / 7.5 | 18515 / 71.6 | 0.9 | 59.3 |
| Niger | 24.2 | 4132 / 170.7 | 138 / 5.7 | 2951 / 71.4 | 3.3 | 61.6 |
| Nigeria | 206.1 | 108943 / 528.6 | 1420/ 6.9 | 85367 / 78.4 | 1.8 | 54.8 |
| Rwanda | 13.0 | 10850 / 834.6 | 140 / 10.8 | 7193 / 66.3 | 1.3 | 68.3 |
| Sénégal | 16.7 | 22738 / 1361.6 | 509 / 30.5 | 19052 / 83.8 | 2.2 | 67.4 |
| Sierra Leone | 8.0 | 2970 / 371.2 | 77 / 9.6 | 2025 / 68.2 | 2.6 | 53.9 |
| South Sudan | 11.2 | 3670 / 327.7 | 63 / 5.6 | 3181 / 86.7 | 1.7 | 57.4 |
| Sudan | 43.8 | 23730 / 541.8 | 1576 / 35.6 | 15240 / 64.2 | 6.6 | 64.9 |
| Togo | 8.3 | 4272 / 514.7 | 73 / 8.8 | 3763/ 88.1 | 1.7 | 60.5 |
| Uganda | 45.2 | 38085 / 842.6 | 304 / 6.7 | 13083 / 34.3 | 0.8 | 62.5 |
| Tanzania | 59.7 | 509 /8.5 | 21 /14.4 | 183/40.0 | 4.1 | 64.5 |
| Mean: SD |  | 926.4 ± 926.2 | 14.4 ± 13.2 | 78.1 ± 16.6 | 2.1 ± 1.4 | 60.7 |

**Table 2.** Non-Ivermectin 22 countries.

| Country | Pop<br>(M) | Morbidity<br>(N/M) | Mortality<br>(N/M) | Recovery<br>(N/%) | Fatality<br>(%) | Life<br>(y/o) |
| --- | --- | --- | --- | --- | --- | --- |
| Algeria | 43.8 | 103611 / 2365.5 | 2831 / 64.6 | 70373 / 67.9 | 2.7 | 76.5 |
| Botswana | 2.3 | 17365 / 7550.0 | 71 / 30.9 | 13519/ 77.8 | 0.4 | 68.8 |
| Cabo Verde | 0.6 | 12901 / 21501.7 | 119 / 198.3 | 12134/ 94.1 | 0.9 | 72.8 |
| Comoros | 0.9 | 1577 / 1752.2 | 41/ 45.6 | 1069 / 67.8 | 2.6 | 63.9 |
| Eritrea | 3.5 | 1877 / 536.3 | 6 / 1.6 | 1073 / 57.2 | 0.3 | 65.5 |
| Eswatini | 1.2 | 12736 / 10613.3 | 360/ 300.0 | 8076 / 63.4 | 2.8 | 58.3 |
| Egypt | 100.0 | 155507 / 155.1 | 8527 / 8.5 | 122291/ 78.6 | 5.5 | 71.8 |
| Gambia | 2.4 | 3897 / 1623.7 | 127/ 52.9 | 3689 / 94.7 | 3.3 | 61.7 |
| Lesotho | 2.1 | 6371 / 3033.8 | 93 / 44.3 | 3022 / 47.4 | 1.5 | 52.9 |
| Libya | 6.9 | 108017 / 15654.6 | 1651 / 239.3 | 85068 / 78.7 | 1.5 | 72.5 |
| Madagascar | 27.7 | 18001 / 649.9 | 267 / 9.6 | 17447 / 96.9 | 1.5 | 66.3 |
| Mauritania | 4.6 | 15999 / 3478.0 | 401 / 87.2 | 14431 / 90.2 | 2.5 | 64.5 |
| Mauritius | 1.3 | 547 / 420.8 | 10 / 7.7 | 516 / 94.3 | 1.8 | 74.5 |
| Morocco | 36.9 | 458865 / 12435.4 | 7911 / 214.4 | 433937 / 94.6 | 1.7 | 76.2 |
| Namibia | 2.5 | 30198 / 12079.2 | 280/ 112.0 | 26458 / 87.6 | 0.9 | 63.4 |
| Sao Tome &<br>Principe | 0.2 | 1130 / 5650 | 17 / 85.0 | 993 / 87.9 | 1.5 | 70.2 |
| South Africa | 59.3 | 1325659 /22355.1 | 36851 / 621.4 | 1083978 / 81.8 | 2.8 | 63.5 |
| Somalia | 15.9 | 4744 / 298.4 | 130 / 8.2 | 3666 / 77.3 | 2.7 | 52.8 |
| Seychelles | 0.1 | 148 / 1480 | 0 / 0 | 147 / 99.3 | 0 | 74.3 |
| Tunisia | 11.8 | 177231 / 15019.6 | 5616 / 475.9 | 127854 / 72.1 | 3.2 | 76.5 |
| Zambia | 18.4 | 36074 / 1960.5 | 537/ 29.2 | 25106 / 69.6 | 1.5 | 63.5 |
| Zimbabwe | 14.9 | 26881 / 1804.1 | 683 / 45.8 | 15872 / 59.0 | 2.5 | 61.2 |
| Mean ± SD |  | 6473.5 ± 7090.5 | 121.9 ±161.2 | 78.0 ± 14.6 | 2.0 ± 1.2 | 66.4 |

### Analytical method

This study aimed to verify the ivermectin intervention statistics in the community-directed treatment with ivermectin. Therefore, we compare the morbidity, mortality, recovery rate, and fatality rate caused by COVID-19 between the two group countries.

### Statistical Analysis

Statistical analysis was done by Microsoft Excel 2016 (Microsoft Corporation Redmond, Washington). Data were presented as a mean and standard deviation (SD) and compared between the two groups utilizing Welch-test. A two-sided P value < 0.05 was considered significant.

## Results

The results of the onchocerciasis-endemic of 31 countries (**Ivermectin group**) and the non-endemic 22 countries (**non-Ivermectin group**) were shown in **Table 3**.

**Table 3.**
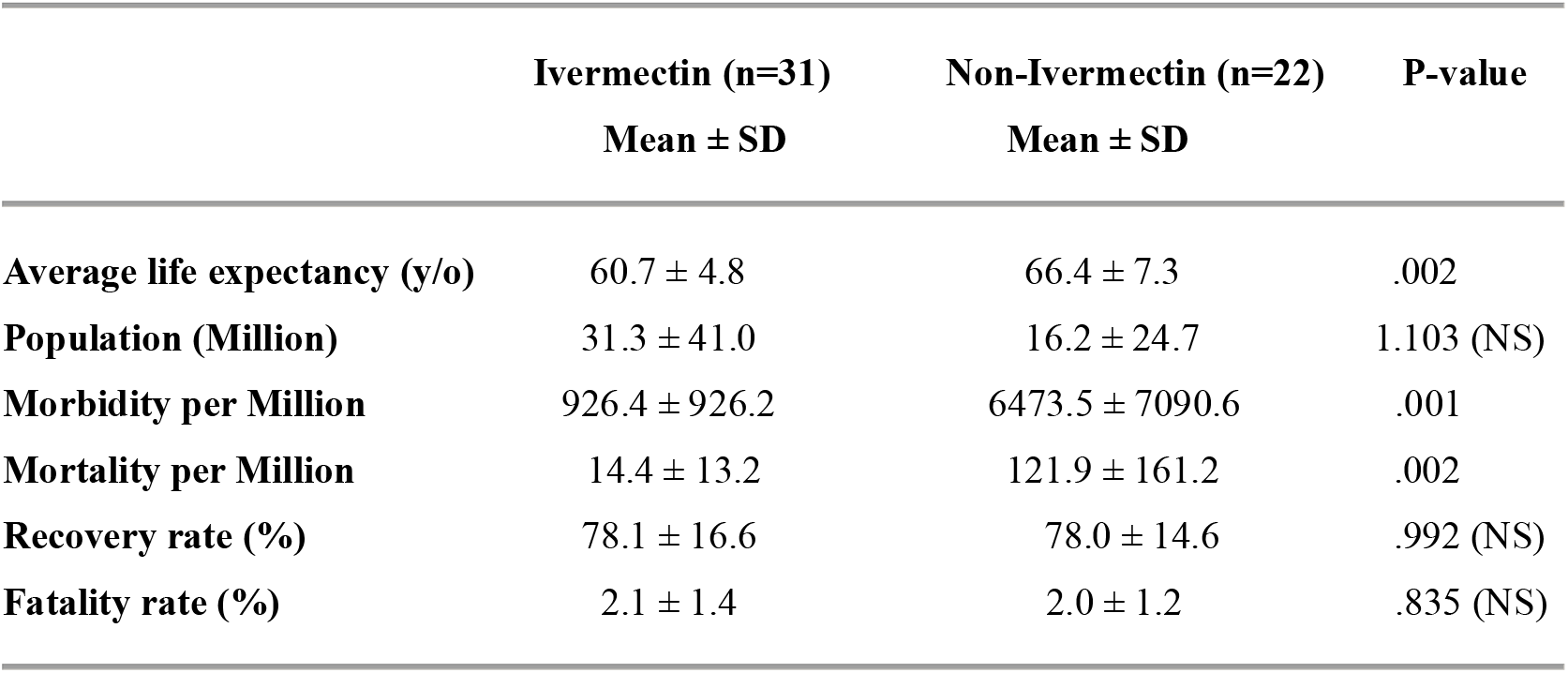
Comparison between the two studied groups.

|  | **Ivermectin (n=31)** | **Non-Ivermectin (n=22)** | **P-value** |
| --- | --- | --- | --- |
|  | **Mean ± SD** | **Mean ± SD** |  |
| **Average life expectancy (y/o)** | 60.7 ± 4.8 | 66.4 ± 7.3 | .002 |
| **Population (Million)** | 31.3 ± 41.0 | 16.2 ± 24.7 | 1.103 (NS) |
| **Morbidity per Million** | 926.4 ± 926.2 | 6473.5 ± 7090.6 | .001 |
| **Mortality per Million** | 14.4 ± 13.2 | 121.9 ± 161.2 | .002 |
| **Recovery rate (%)** | 78.1 ± 16.6 | 78.0 ± 14.6 | .992 (NS) |
| **Fatality rate (%)** | 2.1 ± 1.4 | 2.0 ± 1.2 | .835 (NS) |

Morbidity and mortality were statistically significantly less in the ivermectin group. The population, recovery rate, and fatality rate were not statistically significant differences between them. The average life expectancy was statistically significantly high in the non-ivermectin group.

## Discussion

It is important to verify the effect of ivermectin interventions on the variations in morbidity and mortality and the associated viral lethality. Ivermectin is an approved antiparasitic drug that is used to treat several neglected tropical diseases, including onchocerciasis, helminthiases, and scabies. For these indications, ivermectin has been widely used and has demonstrated an excellent safety profile [10].

This epidemiological study in Africa reveals that the morbidity and mortality in the onchocerciasis-endemic countries (**Ivermectin group**) are statistically lesser than those in the non-endemic ones (**non-Ivermectin group**). However, the morbidity will depend on the number of COVID-19 tests. Onchocerciasis-endemic countries are mainly in the medically weak areas in Sub-Saharan Africa rather than the non-endemic countries. Therefore, the average life is statistically different between the two groups. And, we suggest that the number of COVID-19 tests in the endemic-countries will be lesser than the non-endemic ones because their health systems are under-resourced. In other words, the apparent incidence of COVID-19 may be lower in the onchocerciasis-endemic countries, but mortality is unrelated to the number of tests. The mortality is less in the ivermectin group than the non-ivermectin one. The recovery and fatality rates are statistically the same in both groups, although they will depend on the medical circumstances. That is, is the medical situation not that different between the two group countries, or is there another factor at play?

Some researchers say that the short life expectancy and the fact that 3% of the population is over 65 years old may be a reason for COVID-19 not exploding infections in Africa [1, 2]. The mean age of the average life expectancy in epidemic countries is 60.7 years and 66.4 years in non-epidemic countries. However, countries with shorter average life expectancy have worse medical circumstances than countries with longer average life expectancy. As a result, the epidemic countries have higher infant mortality and lower average life expectancy [8]. And more, if this difference is one of the factors in the impact, the statistically no difference in the recovery rate and fatality rate between the two groups shows a discrepancy. Taking this into account, mortality is lower in ivermectin-treated countries than those in non-treated ones. The recovery rate and fatality rate may be improved according to the number of COVID-19 tests. These results imply that ivermectin will act on SARS-CoV-2, and prevent deterioration in patients. Our results will support those of Mohammad et al.’s study [11].

On the other hand, the South African Health Products Regulatory Authority (SAHPRA) has given the green light for the controlled use of Ivermectin for humans on January 27, 2021 [12]. Since then, the number of new COVID-19 infections in South Africa has been declining.

In conclusion, in the countries where ivermectin is distributed to and used by the entire areas, it suggests to lead a reduction in mortality, to accelerate patient recovery and, to avoid death. And this analytical study will suggest that early treatment with ivermectin may accelerate recovery and prevent worsening of symptoms in patients with mild disease. These findings can be efficiently translated into therapies for SARS-CoV-2 (COVID-19).

## Data Availability

All data is publicly available.

https://unstats.un.org/unsd/demographic-social/products/vitstats/seratab2.pdf

https://www.who.int/publications/m/item/weekly-epidemiological-update---19-january-2021

## Ethical approval

Ethical approval was not required for this work as public data analysis.

## Authors’ contributions

H.T., S.T., and K.K. conceived and designed the study. H.T. and S.T. analyzed the data and wrote the first draft. All authors provided critical revisions. All authors contributed equally to this study and approved the submitted manuscript.

